# REAL-WORLD EFFECTIVENESS OF NIRMATRELVIR/RITONAVIR ON COVID-19-ASSOCIATED HOSPITALIZATION PREVENTION: A POPULATION-BASED COHORT STUDY IN THE PROVINCE OF QUÉBEC, CANADA

**DOI:** 10.1101/2023.02.14.23285860

**Authors:** J.L. Kabore, B. Laffont, M. Diop, M.R. Tardif, A. F. Turgeon, J. Dumaresq, M. Luong, M. Cauchon, H. Chapdelaine, D. Claveau, M. Brosseau, E. Haddad, M. Benigeri

## Abstract

**Introduction:** The nirmatrelvir/ritonavir (PAXLOVID™) is an antiviral blocking the replication of SARS-CoV-2. Early treatment with this antiviral has showed to reduce COVID-19 hospitalization and death in unvaccinated outpatients with mild-to-moderate COVID-19 and high risk of progression to severe disease with variants before Omicron. However, the current epidemiological context and the level of immunity in the population (vaccination and/or natural infection) have evolved considerably since the disclosure of these results. Thus, real-world evidence studies in vaccinated outpatients with lineage and sublineage of the variant are needed.

**Objective:** To assess whether nirmatrelvir/ritonavir treatment reduces the risk of COVID-19-associated hospitalization among Québec outpatients with mild-to-moderate COVID-19 at high risk of progression to severe disease in a real-world context, regardless of vaccination status and circulating variants, in the province of Québec.

**Methods:** This was a retrospective cohort study of SARS-CoV-2-infected outpatients who received nirmatrelvir/ritonavir between March 15 and August 15, 2022, using data from the Québec provincial clinico-administrative databases. Outpatients treated with nirmatrelvir/ritonavir were compared to unexposed ones. The treatment group was matched with controls using propensity-score matching in a ratio of 1:1. The outcome was COVID-19-associated hospitalization occurring within 30 days following the index date. Poisson regression with robust error variance was used to estimate the relative risk of hospitalization among the treatment group compared to the control group.

**Results:** A total of 16,601 and 242,341 outpatients were eligible to be included in the treatment (nirmatrelvir/ritonavir) and control groups respectively. Among treated outpatients, 8,402 were matched to controls. Regardless of vaccination status, nirmatrelvir/ritonavir-treated outpatient status was associated with a 69% reduced relative risk of COVID-19-associated hospitalization (RR: 0.31 [95% CI: 0.28; 0.36]). The effect was more pronounced in outpatients without a complete primary vaccination course (RR: 0.04 [95% CI: 0.03; 0.06]), while treatment with nirmatrelvir/ritonavir was not associated with benefit when outpatients with a complete primary vaccination course were considered (RR: 0.93 [95% CI: 0.78; 1.08]) Subgroups analysis among outpatients with a primary vaccination course showed that nirmatrelvir/ritonavir treatment was associated with a significant decrease in relative risk of hospitalization in severely immunocompromised outpatients (RR: 0.66 [95% CI: 0.50; 0.89]) and in outpatients aged 70 years and older (RR: 0.50 [95% CI: 0.34; 0.74]) when the last dose of the vaccine was received more than six months before.

**Conclusions:** Among SARS-CoV-2-infected outpatients at high risk for severe COVID-19 during Omicron BA.2 and BA.4/5 surges, treatment with nirmatrelvir/ritonavir was associated with a significant reduced relative risk of COVID-19-associated hospitalization. This effect was observed in outpatients with incomplete primary vaccination course and in outpatients who were severely immunocompromised. Except for severely immunocompromised outpatients, no evidence of benefit was found in any category of outpatient with a complete primary vaccination course whose last dose of COVID-19 vaccine was received within six months.

## INTRODUCTION

The coronavirus disease 2019 (COVID-19) pandemic, caused by severe acute respiratory syndrome coronavirus 2 (SARS-CoV-2), has been one of the greatest threats to public health in the 21st century, with more than 755 million identified cases and over 6.8 million deaths reported worldwide (as of February 13, 2023) ^1^.

Since the beginning of the pandemic, several interventions were tested but few targeted non-hospitalized. Among those, the combination of nirmatrelvir/ritonavir was proven potentially beneficial. Following the United States Food and Drug Administration (FDA) Emergency Use Authorization (EUA) in December 2021, Health Canada (HC) issued, in January 2022, a notice of compliance for early treatment with nirmatrelvir/ritonavir in persons with mild-to-moderate cases of COVID-19 who are at high risk for progression to severe disease. The primary data supporting the HC authorization came from the EPIC-HR trial, performed during an infection wave driven by the Delta variant. Furthermore, all participants in EPIC-HR were unvaccinated for COVID-19 and did not have a documented SARS-CoV-2 infection. EPIC-HR showed a risk reduction of 89% for hospitalization or death in patients treated with nirmatrelvir/ritonavir, when treatment was provided for five consecutive days, as soon as possible after a confirmed SARS-CoV-2 infection and within five days of symptom onset ^2^.

However, the emergence of the fast-spreading SARS-CoV-2 Omicron variant in late 2021 led to a global COVID-19 resurgence. By the end of December 2021, Omicron was outcompeting the Delta variant, and by early January 2022, Omicron had spread widely worldwide, even in countries with high levels of pre-existing immunity ^3,4^. As of today, Québec outpatients eligible for treatment with nirmatrelvir/ritonavir are those with severe immunosuppression (regardless of their vaccination status), as well as those without a complete primary vaccination course who are aged 60 and over or who are aged 18 and over and have at least one risk factor for severe COVID-19. Outpatients with a complete primary vaccination course who are at high risk of severe COVID-19 are also eligible for nirmatrelvir/ritonavir on a case-by-case basis, as judged by the clinician ^5^.

As the effectiveness of nirmatrelvir/ritonavir against the Omicron variants and sub-variants as well as in outpatients with prior COVID-19 immunity remains uncertain, we designed this large provincial population-based study to assess, in a real-world context, the effectiveness of nirmatrelvir/ritonavir therapy for preventing COVID-19-associated hospitalizations in high-risk Québec outpatients during a BA.2 and BA.4/5 Omicron surge.

## METHODS

### Setting and data

This was a population-based retrospective cohort study of patients treated with nirmatrelvir/ritonavir between March 15, and October 15, 2022, in the province of Québec, Canada.

Data came from the provincial Québec Public Health Insurance (Régie de l’assurance maladie du Québec [RAMQ databases]) and from the Ministry of Health and Social Services (Ministère de la Santé et des Services sociaux [MSSS databases]). RAMQ databases included sociodemographic characteristics of insured patients (age, sex, date of death, etc.), information on drug dispensation (drug identification number (DIN) and non-proprietary name, dosage, treatment duration, etc.) and information on medical visits (date, diagnosis encoded ICD-10, physician specialties, etc.). MSSS databases included data on emergency visits and hospitalizations (admission and discharge dates, encoded ICD-10 diagnosis, etc.) and data on COVID-19. The COVID-19 database included date of infection (positive RT-PCR test), sociodemographic patient characteristics (age, sex, region of residence, etc.) and information on the occurrence of COVID-19-associated hospitalization, and COVID-19-associated death. Vaccination information (administration dates for the first and subsequent vaccine doses) was also recorded in COVID-19 databases. A unique, anonymised identifier was used to match individual data from these different databases (RAMQ and MSSS databases).

### Ethical Considerations

Access to these databases was made possible through a tripartite agreement between the MSSS, the RAMQ and the Institut national d’excellence en santé et en services sociaux (INESSS), which has been approved by the Commission d’accès à l’information du Québec.

### Population

The study population included individuals covered by the Québec public health insurance plan in 2022 and was divided into two groups: those who received nirmatrelvir/ritonavir medication (treatment group) and those who did not (control group). The treatment group included outpatients who received at least one prescription of nirmatrelvir/ritonavir between March 15, and October 15, 2022, in community pharmacies. Prescriptions of nirmatrelvir/ritonavir were identified through the DIN codes. The control group included outpatients who tested positive for COVID-19 (RT-PCR test) between March 15, and October 15, 2022, but who did not receive a prescription of nirmatrelvir/ritonavir during the study period. For the control group with several positive RT-PCR tests during this period, only the first positive test date was considered. All included outpatients were followed until November 14, 2022. The index date was the start date of the follow-up period and was defined as the date of the first prescription of nirmatrelvir/ritonavir for the treatment group, or as the date of the COVID-19 positive test for the control group. As of January 2022, RT-PCR testing was restricted to priority groups which included healthcare and social service workers, healthcare and social service clients, and other high-risk individuals (individuals aged 70 years and older, individuals under consideration for COVID-19 or for flu treatment, individuals with physical or intellectual disability, etc.) Thus, most of the individuals who received nirmatrelvir/ritonavir prescription did not have a positive RT-PCR test recorded in the study databases. Occurrence of hospitalization in the 30 days following the index date was recorded. Individuals with less than 30 days of follow-up were excluded. Patients already hospitalized at the time of the COVID-19 positive test were excluded. Patients living in long-term care facilities were also excluded, because they are likely to be treated in long-term care facilities and this information was not available in the databases (Figure 1).

**Figure 1.**
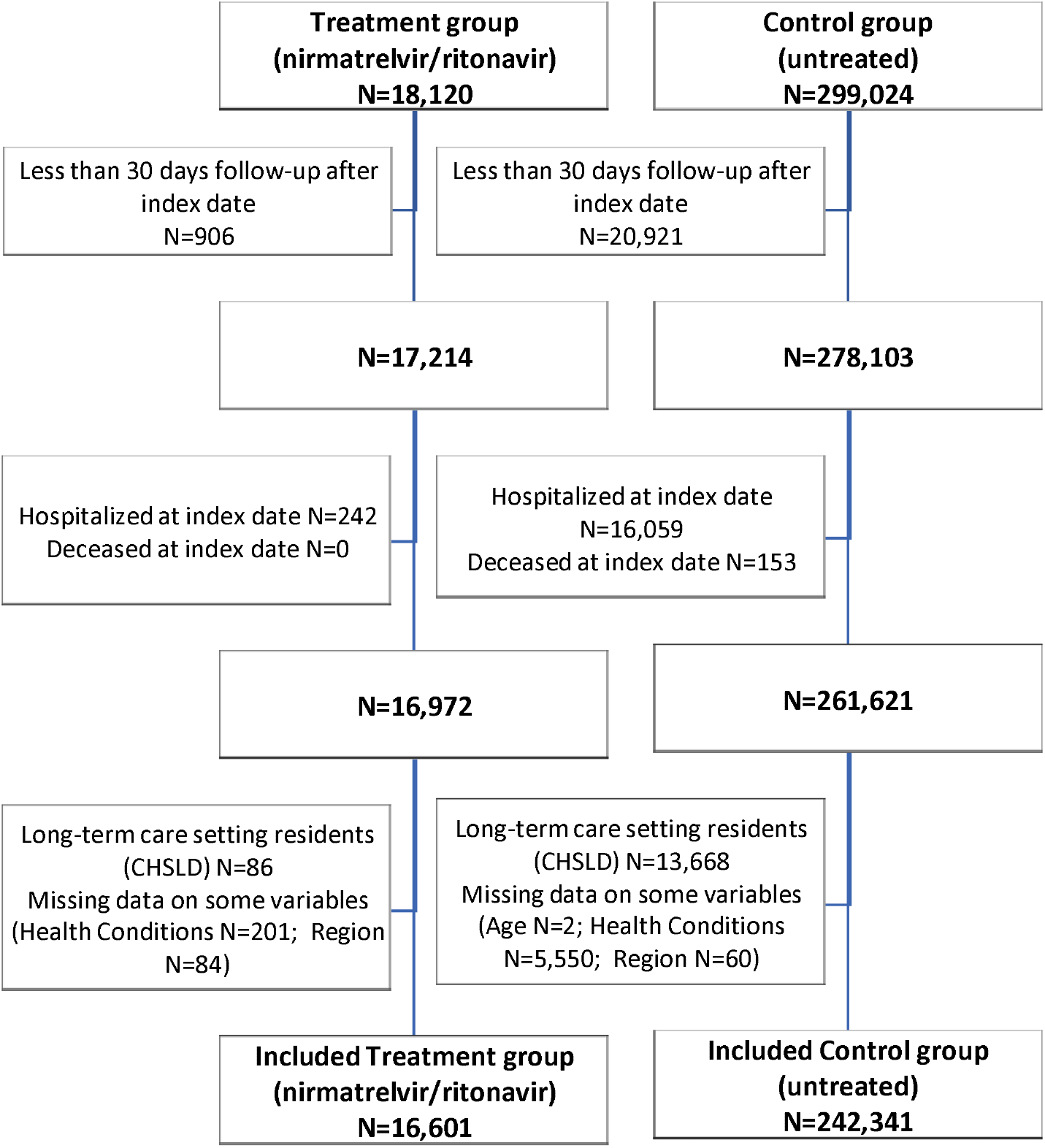
Flow chart of outpatients (treatment and control groups) inclusion and exclusion

### Variables

Variables were determined or calculated at the index date and comprised socio-demographic characteristics, COVID-19 data, and health conditions. The sociodemographic characteristics included age, sex, and regions of residence. COVID-19 data included vaccination status (with complete primary vaccination course or without complete primary vaccination course), number of vaccine doses, time since the last vaccine dose, and COVID-19 epidemic waves. The first dose is considered effective 14 days after administration, whereas subsequent doses (two doses or more) are considered effective after 7 days. Individuals with a complete primary vaccination course included those who received at least two vaccine doses (as opposed to individuals who received no vaccine or only one vaccine dose). This definition of complete primary vaccination (2 doses) was also applied to severely immunocompromised individuals. Time since the last dose of the vaccine was categorized into less than 3 months, 3 to 6 months, 6 to 9 months, 9 to 12 months, and more than 12 months. Individuals with a complete primary vaccination course were divided into 2 groups according to the time since the last vaccine dose: less than 6 months (≤ 182 days) and more than 6 months (>182 days).

Health conditions were identified through Population Grouping Methodology (POP Grouper), developed by the Canadian Institute for Health Information (CIHI) ^6^.The POP Grouper is a methodology that uses a case-mix classification to profile each person in the population using clinical information (ICD-9 and ICD-10 diagnosis codes) in the past three years. This algorithm identifies 226 health conditions. The number of health conditions (0, 1 to 4, 5 to 9, 10 to 14, 15 to 19 and 20 or more) and types of health conditions (respiratory and cardiovascular conditions, immunosuppression conditions and cancer conditions) were considered in this analysis. Details on diagnosis included in each type of health condition were provided in Appendix D.

For the purposes of subgroup analysis of individuals at higher risk of COVID-19-related complications, those with severe immunosuppression were identified. Severe immunosuppression was defined as receiving high-dose immunosuppressive drugs (immunosuppressive drugs in solid organ transplants, anti-cell B therapy, alkylating agents, systemic corticosteroids) with a treatment duration which encompassed the index date or having received a haematological cancer diagnosis (leukaemia, lymphoma, multiple myeloma) in the 12 months prior to the index date. Details about severe immunosuppression are provided in Appendix D.

### Outcome

The outcome was the COVID-19-associated hospitalization in the 30 days following the index date, which included hospitalization with COVID-19 (positive test at admission but not recorded as the main cause of admission) and hospitalization due to COVID-19 (recorded as the main cause of admission). The hospitalization occurring in the 30 days following the index date was considered.

### Statistical Analysis

Descriptive statistics were used to characterize the population that received at least one prescription for nirmatrelvir/ritonavir and to characterize the proportion of cases hospitalized within 30 days of the date of the first prescription for nirmatrelvir/ritonavir. For the purposes of regression analyses, individuals with missing data were excluded, as they represent a small number.

The relative risk of hospitalization in the treatment group compared to the control group was estimated using a Poisson regression with robust error variance. This modified Poisson regression procedure is more reliable than binomial regression and at least as flexible and powerful ^7^. Propensity score matching was performed to reduce confounding and to ensure that the treatment group and the control group were comparable. Propensity score was performed through a logistic regression with the nirmatrelvir/ritonavir prescription as the dependent variable (Yes/No) and the confounding variables as independent variables (age, sex, region of residence, number of vaccine doses, time since the last vaccine dose, COVID-19 epidemic waves, number of health conditions, respiratory and cardiovascular condition, immunosuppression condition, cancer condition). Each individual from the treatment group was then matched to an individual in the control group using the one-to-one nearest neighbour matching method with a calliper of 10^−5^. The calliper was reduced to 10^−2^ for severely immunocompromised individuals to obtain sufficient matches and to increase statistical power for the regression model. The matching was carried out without replacement, i.e., each individual from the control group was used once to be matched with one individual in the treatment group. Poisson regression with robust error variance was then used to estimate the relative risk of hospitalization in the treatment group compared to the control group. Confounding variables used in the propensity score were included again in the final regression model to further minimize the differences between the matched groups. Propensity score matching was repeated for each subgroup analysis. Subgroups analyses included analysis in individuals without complete primary vaccination course, with complete primary vaccination course (overall), according to age (less than 70 years and 70 years and older), and with severe immunosuppression.

Using raw data on hospitalization, we calculated the number needed to treat (NNT) with a 95% confidence interval, when relevant. This calculation was made for all individuals. The relative risk of occurrence of the event was calculated for each population and the NNT was then calculated according to the following formula ^8^:

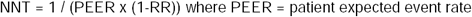

A sensitivity analysis with multivariable regression including overall individuals (not only matches individuals) was also performed to estimate the relative risk of hospitalization (significance threshold used: P-value < .05). Finally, a sensitivity analysis was carried out considering only hospitalizations for which the main cause of admission was COVID-19 as the outcome, as well as another sensitivity analysis considering both COVID-19-associated hospitalization and death as the outcome. Analyses were performed using Stata/SE 15.1 for Windows (StataCorp LLC).

## RESULTS

### Study population

A total of 16,601 and 242,341 outpatients were included in treatment and control groups respectively. Outpatients were included if they had a COVID-19 diagnosis between March 15, and October 15, 2022, and were followed until November 14, 2022, to ensure 30 days of follow-up for each patient. A flow chart depicting study population selection for the analyses on the relative risk of hospitalization is shown in Figure 1. The baseline sociodemographic and clinical characteristics of eligible outpatients are shown in Table 1.

**Table 1.**
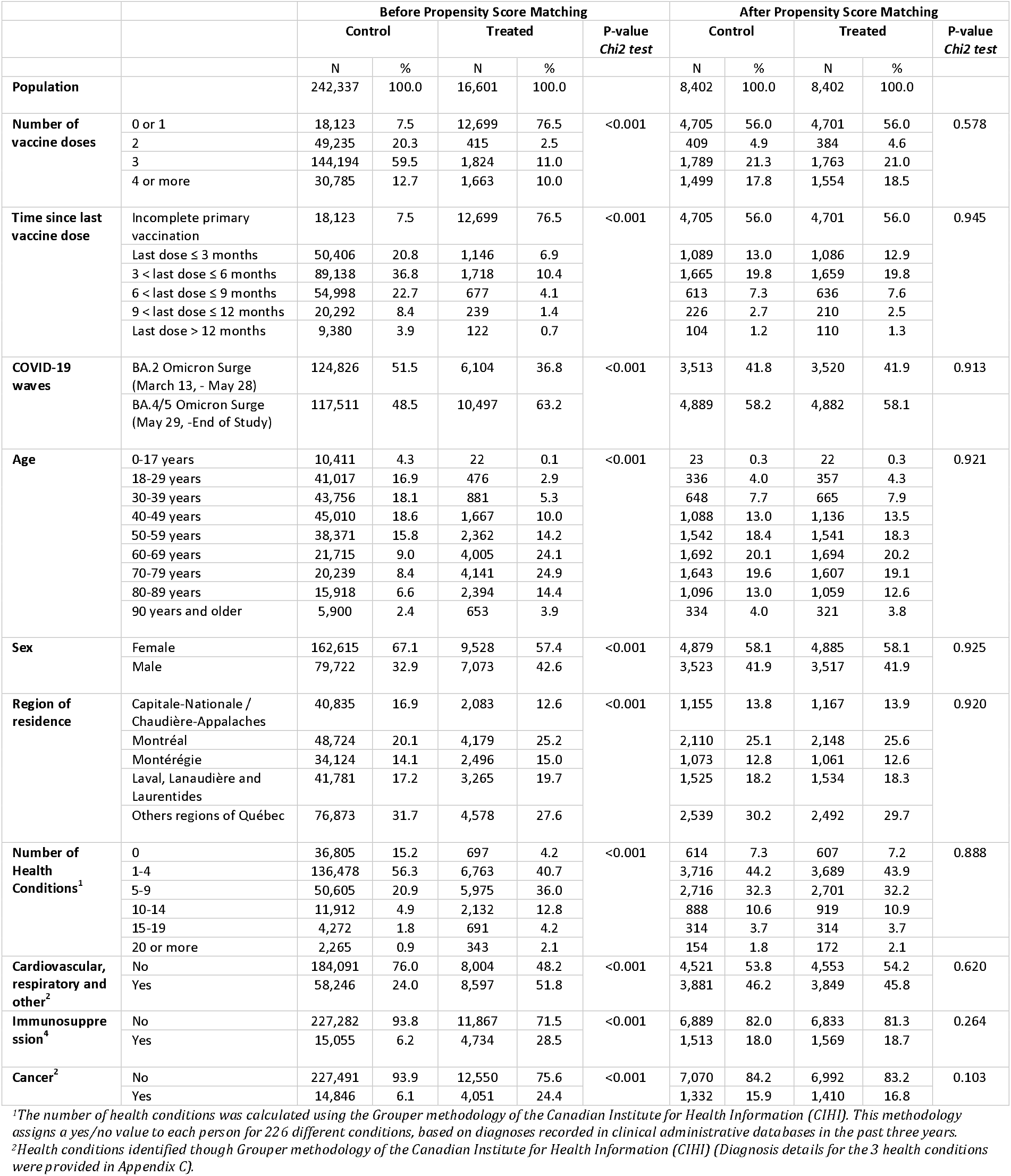
Outpatients’ characteristics before and after propensity score matching and between treatment and control group

### Outpatients Treated With Nirmatrelvir/Ritonavir

A total of 16,601 outpatients received nirmatrelvir/ritonavir in the province of Québec during the study period. Among them, 57% were women, 67% were 60 years or older and 76% did not complete their primary vaccination course, while 73% had at least one targeted health conditions at high risk of complications (respiratory and cardiovascular [52%], immunosuppression [29%] and cancer [24%]). Positive RT-PCR test was reported in 2% of outpatients who did not complete primary vaccination course and in 63% of those who completed primary vaccination course. The baseline sociodemographic and clinical characteristics of outpatients treated with nirmatrelvir/ritonavir are shown in Table 2. Among outpatients who received nirmatrelvir/ritonavir, 2.1% were subsequently hospitalized, with the highest rates of hospitalization observed among older outpatients: 4.3% of those aged 80–89 and 6.9% of those aged 90 years and over (Table 3).

**Table 2.**
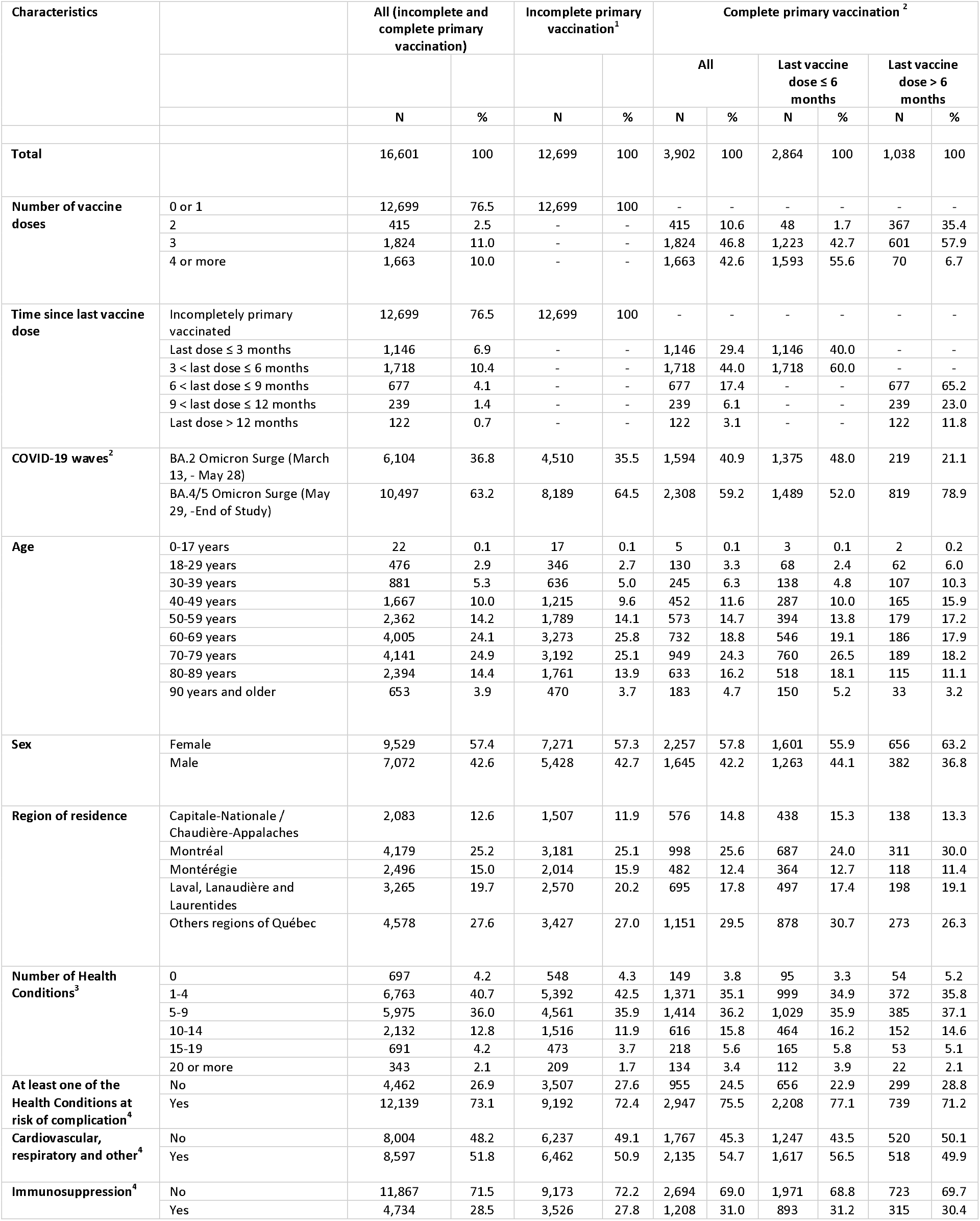
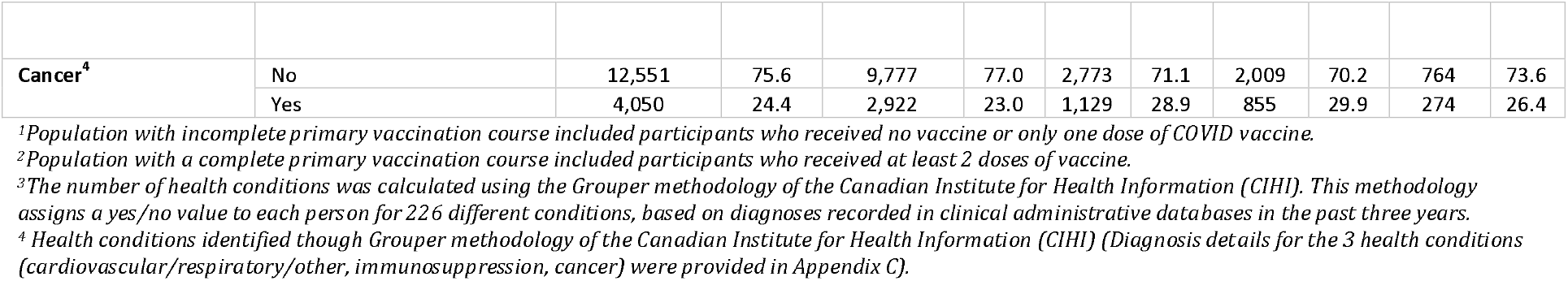
Characteristics of outpatients who received nirmatrelvir/ritonavir prescription (treatment group)

**Table 3.**
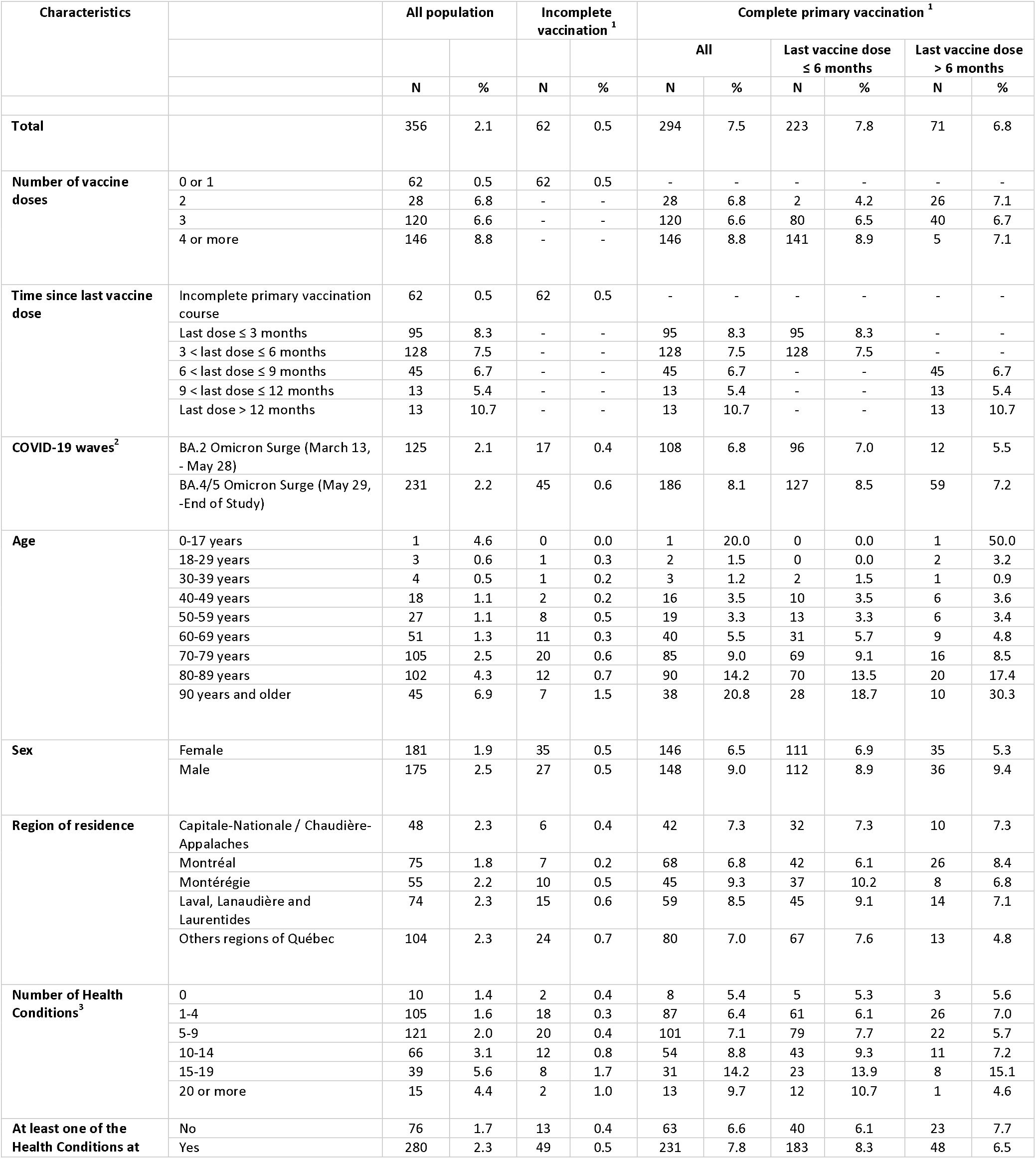
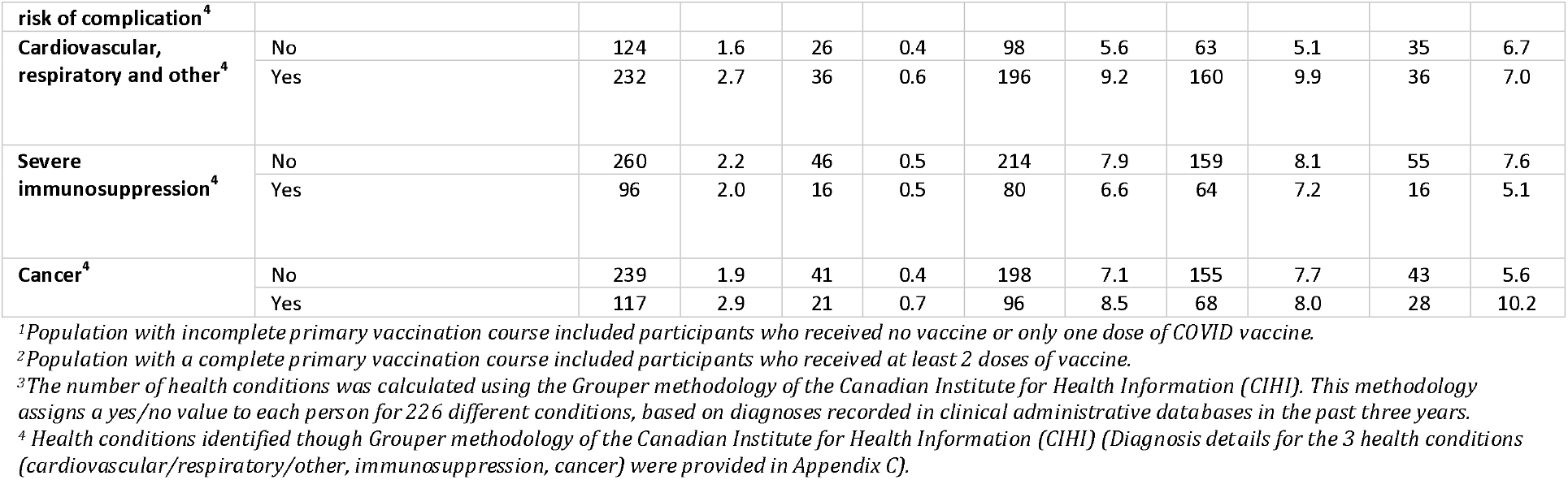
Number and percentage of hospitalizations occurring in the 30 days following index date among nirmatrelvir/ritonavir group (treatment group) and according to the vaccination status

The propensity score matching left 16,804 outpatients (8,402 in each group) with 58% women, 57% aged 60 years or older and 56% without complete primary vaccination course. In addition, 51% had five or more comorbidities, 46% had cardiovascular or respiratory disease, 18% were severely immunocompromised and 16% had cancer.

### Effectiveness of Nirmatrelvir/Ritonavir

Regardless of vaccination status, treatment with nirmatrelvir/ritonavir was associated with a 69% reduction of relative risk of COVID-19-associated hospitalization among infected outpatients with high risk of complications (RR: 0.31 [95% CI: 0.28; 0.36]; p < .001) (Table 4). The NNT with nirmatrelvir/ritonavir to obtain one less event than in the control group was 13 (NNT: 13 [95% CI: 12; 14]). The effect was observed to be more important in those with an incomplete primary vaccination course (RR: 0.04 [95% CI: 0.03; 0.06]; p < .001; NNT: 8 [95% CI: 8; 8]) with a 96% reduced relative risk (Table 4). However, treatment with the antiviral had no significant effect in outpatients with a complete primary vaccination course (RR: 0.93 [95% CI: 0.78; 1.08]; p = .321) (Table 4). Results are comparable in multivariate analyses (Appendix A, Table 1).

**Table 4.**
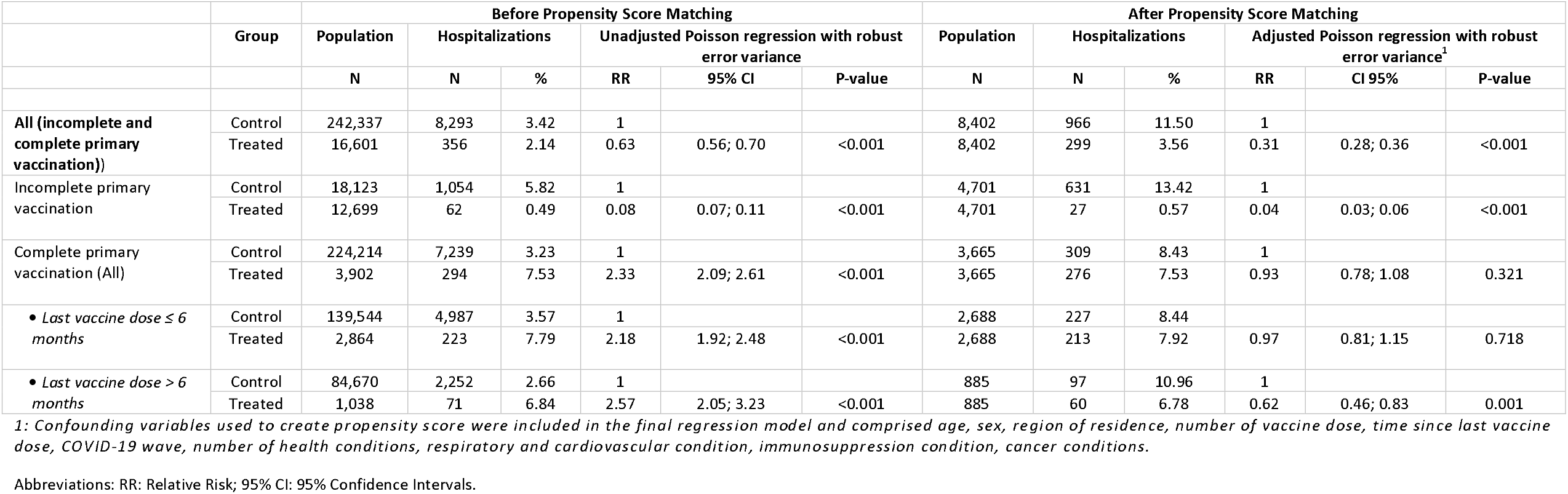
Risk of hospitalization among outpatient with high-risk of progression to severe COVID-19 who received nirmatrelvir/ritonavir prescription compared to controls

In high-risk outpatients with a complete primary vaccination course, the time since the last dose of the vaccine and age impacted the results. No significant effect of nirmatrelvir/ritonavir was observed on the risk of hospitalization in those whose last dose was received within six months, independent of age (RR: 0.97 [95% CI : 0.81; 1.15]; p = .718; Table 4), while a 38% reduced relative risk of COVID-19-associated hospitalization was observed in those whose last dose was received more than six months before (RR: 0.62 [95% CI : 0.46; 0.83]; p = .001; NNT: 24 [95% CI : 17; 58]; Table 4). Treatment with nirmatrelvir/ritonavir was associated with a 25% reduced relative risk of COVID-19-associated hospitalization in high-risk outpatients aged 70 and older (RR: 0.75 [CI 95%: 0.63; 0.88]; p = .001; NNT: 28 [95% CI: 18; 68]) (Table 5). The effect was even stronger in those aged 70 and older whose last dose was received more than six months before (RR: 0.50 [CI 95%: 0.34; 0.74]; p < .001; NNT: 10 [95% CI: 7; 20]) (Table 5), whereas no significant effect was observed in outpatients aged 70 and over when the last dose of the vaccine was received six months before or less (RR: 0.89 [95% CI : 0.73; 1.08]; p = .240) (Table 5). Importantly, use of nirmatrelvir/ritonavir has no effect on COVID-19-associated hospitalization for completely primary vaccinated outpatients aged less than 70 years, regardless of the time elapsed since the last dose of vaccine (Table 5).

**Table 5.** Subgroup analyses of outpatients with high-risk of progression to severe COVID-19 and complete primary vaccination on the risk of hospitalization in participants who received nirmatrelvir/ritonavir prescription compared to controls

| Population | Group | Before Propensity Score Matching |  |  |  |  |  | After Propensity Score Matching |  |  |  |  |  |
| --- | --- | --- | --- | --- | --- | --- | --- | --- | --- | --- | --- | --- | --- |
|  |  | Population | Hospitalization |  | Unadjusted Poisson regression with robust error variance |  |  | Population | Hospitalization |  | Adjusted Poisson regression with robust error variance<sup>1</sup> |  |  |
|  |  | N | N | % | RR | 95% CI | P-value | N | N | % | RR | 95% CI | P-value |
| Less than 70 years (All) | Control | 183,690 | 2,037 | 1.1 | 1 |  |  | 1,869 | 63 | 3.4 | 1 |  |  |
|  | Treated | 2,137 | 81 | 3.8 | 3.42 | 2.75; 4.25 | <0.001 | 1,869 | 74 | 4.0 | 1.20 | 0.87; 1.65 | 0.269 |
| Less than 70 years (last dose ≤ 6 months) | Control | 105,265 | 1,136 | 1.1 | 1 |  |  | 1,231 | 40 | 3.3 | 1 |  |  |
|  | Treated | 1,436 | 56 | 3.9 | 3.61 | 2.78; 4.70 | <0.001 | 1,231 | 48 | 3.9 | 1.20 | 0.81; 1.81 | 0.361 |
| Less than 70 years (last dose > 6 months) | Control | 78,425 | 901 | 1.2 | 1 |  |  | 579 | 16 | 2.8 | 1 |  |  |
|  | Treated | 701 | 25 | 3.6 | 3.10 | 2.10; 4.59 | <0.001 | 579 | 20 | 3.5 | 1.31 | 0.68; 2.49 | 0.418 |
| 70 years and older (All) | Control | 40,524 | 5,202 | 12.8 | 1 |  |  | 1,678 | 261 | 15.6 | 1 |  |  |
|  | Treated | 1,765 | 213 | 12.1 | 0.94 | 0.83; 1.07 | 0.346 | 1,678 | 199 | 11.9 | 0.75 | 0.63; 0.88 | 0.001 |
| 70 years and older (Last dose ≤ 6 months) | Control | 34,279 | 3,851 | 11.2 | 1 |  |  | 1,388 | 181 | 13.0 | 1 |  |  |
|  | Treated | 1,428 | 167 | 11.7 | 1.04 | 0.90; 1.20 | 0.589 | 1,388 | 161 | 11.6 | 0.89 | 0.73; 1.08 | 0.240 |
| 70 years and older (Last dose > 6 months) | Control | 6,245 | 1,351 | 21.6 | 1 |  |  | 253 | 58 | 22.9 | 1 |  |  |
|  | Treated | 337 | 46 | 13.7 | 0.63 | 0.48; 0.83 | 0.001 | 253 | 30 | 11.9 | 0.50 | 0.34; 0.74 | <0.001 |
| Severely immunocompromised (All) | Control | 2,012 | 452 | 22.5 | 1 |  |  | 519 | 95 | 18.3 | 1 |  |  |
|  | Treated | 524 | 63 | 12.0 | 0.54 | 0.42; 0.68 | <0.001 | 519 | 62 | 12.0 | 0.66 | 0.50; 0.89 | 0.005 |
| Severely immunocompromised (Last dose ≤ 6 months) | Control | 1,502 | 334 | 22.2 | 1 |  |  | 369 | 78 | 21.1 | 1 |  |  |
|  | Treated | 375 | 44 | 11.7 | 0.53 | 0.39; 0.71 | <0.001 | 369 | 43 | 11.7 | 0.58 | 0.41; 0.81 | 0.001 |
| Severely immunocompromised (Last dose > 6 months) | Control | 510 | 118 | 23.1 | 1 |  |  | 139 | 32 | 23.0 | 1 |  |  |
|  | Treated | 149 | 19 | 12.8 | 0.55 | 0.35; 0.86 | 0.009 | 139 | 19 | 13.7 | 0.61 | 0.37; 0.99 | 0.047 |

1: Confounding variables used to create propensity score were included in the final regression model and comprised age, sex, region of residence, number of vaccine dose, time since last vaccine dose, COVID-19 wave, number of health conditions, respiratory and cardiovascular condition, immunosuppression condition, cancer conditions.
Abbreviations: RR: Relative Risk; 95% CI: 95% Confidence Intervals.

Nirmatrelvir/ritonavir was associated with a 34% reduction of relative risk of COVID-19-associated hospitalization in severely immunocompromised patients with a complete primary vaccination course (RR: 0.66 [95% CI : 0.50; 0.89]; p = .005; NNT: 16 [95% CI : 11; 45]) (Table 5), regardless of the time since their last dose of vaccine (six months or less: RR: 0.58 [95% CI : 0.41 ; 0.81]; p = .001; NNT: 11 [95% CI : 8 ; 22]; more than six months: RR: 0.61 [95% CI : 0.37 ; 0.99]; p = .047; NNT: 11 [95% CI : 7 ; 945]) (Table 5).

Sensitivity analyses focused only on hospitalization due to COVID-19 (i.e., hospitalization with COVID-19 as the main cause of admission) showed comparable results to the primary analysis. Treatment with nirmatrelvir/ritonavir was associated with a significant reduction of relative risk of hospitalization due to COVID-19 in the global population, regardless of vaccination status (Appendix B, Table 1), as well as in outpatients without a complete primary vaccination course (Appendix B, Table 1). As previously observed, for those with a complete primary vaccination course, time since the last dose of vaccine impacted the results, with a reduced relative risk of hospitalization due to COVID-19 in those with last vaccine dose received more than six months before (Appendix B, Table 1), but not in those whose last dose was received within six months.

## DISCUSSION

In this real-world study and province-wide study, the treatment with nirmatrelvir/ritonavir in outpatients at high-risk of severe COVID-19 with incomplete primary vaccination course was shown to be associated with lower risk of COVID-19-associated hospitalization in the province of Québec during a period where BA.2 and the BA.4/5 were the dominant variants. Comparable observations were made in outpatients with high risk of progression to severe disease and a complete primary vaccination course who received their last dose of vaccine more than six months before, a result driven by vaccinated outpatients aged 70 and over. Treatment with nirmatrelvir/ritonavir had no significant effect in outpatients with high risk of progression to severe disease and a complete primary vaccination course less than six months ago, suggesting that an up-to-date vaccination status (booster dose at 6 months) against SARS-CoV-2 remains an effective method for preventing severe illness ^9,10^. A reduced risk of hospitalization was also observed with nirmatrelvir/ritonavir in severely immunocompromised outpatients, regardless of their vaccination status or the time since the last dose of the vaccine. These findings confirm the ability of nirmatrelvir/ritonavir to reduce the risk of hospitalization in individuals with high risk of progression to severe disease and infected with different Omicron variants of SARS-Cov-2.

The observed results in outpatients without a complete primary vaccination course were slightly larger than those observed in the EPIC-HR trial (89%) ^2^. This might be explained by differences related to the study populations, variants, study design and settings. A greater representation of older as well as severely immunocompromised individuals may explain the greater effect observed in this study. Furthermore, in the EPIC-HR trial participants were almost exclusively infected by the delta variant (98%), while our study was performed when Omicron BA.2 and BA.5 were predominant in Québec. It has also been described that Omicron variants have been observed to cause lower rates of severe cases ^11-15^. The results observed in outpatients with a complete primary vaccination course at high risk of severe COVID-19 were in line with the EPIC-SR trial results, where a non-significant reduction in hospitalizations and death was observed in nirmatrelvir/ritonavir-treated vaccinated patients with at least one risk factor for severe COVID-19 ^16^. The subgroup analyses revealed that time since the last dose of the vaccine is important to consider, given the antiviral’s effectiveness outpatients with a complete primary vaccination course and high risk of severe infection. Comparable results were reported in previous study where treatment with nirmatrelvir/ritonavir within 20 weeks of the last dose of the vaccine was not associated with a reduced risk of hospitalization, whereas a significant reduced risk has been reported following the use of nirmatrelvir/ritonavir in infected outpatients whose last dose of the vaccine was received more than 20 weeks before ^17^. These results might be explained by a poorer response to vaccination or by senescence of the immune system in older individuals, since that study was exclusively performed in individuals aged 50 years and older, with 41% aged 65 years and older ^17^. Overall, our data support several observational studies in which the use of nirmatrelvir/ritonavir was associated with a reduced risk of hospitalization or severe COVID-19 in outpatients infected by SARS-CoV-2 at higher of complications ^17-40^.

Data from observational studies carried out during Delta and Omicron surges also provide real-world confirmation of the ability of nirmatrelvir/ritonavir to neutralize the different variants of SARS-CoV-2. Combined with its ease of administration, this makes nirmatrelvir/ritonavir one of the preferred treatment options for infected outpatients at high risk of severe COVID-19, especially since several alternative options are not able to neutralize certain variants of SARS-CoV-2.

However, despite nirmatrelvir/ritonavir’s demonstrated effectiveness and ease of administration, it should be noted that nirmatrelvir/ritonavir is not recommended in several medical conditions and has complex drug-drug interactions due to the ritonavir component of the combination ^41^. Therefore, the use of nirmatrelvir/ritonavir requires particular vigilance in order to review the patient’s medication history, determine potential drug interactions and/or prescribe any required adjustments to concomitant medications. This may limit the use of nirmatrelvir/ritonavir in high-risk patients, for whom other alternative antivirals or monoclonal antibodies might be appropriate ^42-45^.

Clinical data suggest that patients treated with nirmatrelvir/ritonavir may experience a resurgence of COVID-19, although to date these phenomena have only been associated with mild-stage COVID-19 cases ^21,46-59^. A brief return of symptoms may be part of the natural history of SARS-CoV-2 infection, regardless of nirmatrelvir/ritonavir use or vaccination status, and there is no evidence to recommend additional treatment with nirmatrelvir/ritonavir or another therapy for SARS-CoV-2 when a resurgence of COVID-19 is suspected. An observational study carried out in individuals infected with SARS-CoV-2 who received no pharmacological treatment reported that recrudescence of COVID-19 had been observed in 10% of study participants ^60^. Besides, regardless of vaccination status and infection history, another study also reported that treatment with nirmatrelvir/ritonavir, compared to no antivirals or monoclonal antibodies, is associated with a reduced risk of post-infection sequelae in the cardiovascular system (dysrhythmia and ischaemic heart disease), coagulation and haematological disorders (deep vein thrombosis and pulmonary embolism), fatigue, liver disease, acute kidney disease, muscle pain, neurocognitive impairment and shortness of breath ^39^.

Our study has several limitations. As in any observational study, confounding factors such as clinical and sociodemographic characteristics may have biased the observed effectiveness. Confounding factors such as obesity, tobacco use, as well as severity of comorbidities were not available in our databases and thus were not included in propensity score. In addition, severity of symptoms and time interval between onset of symptoms and antiviral prescriptions were not available and may have confounded the results. It should also be noted that there is a lack of information on the actual use of nirmatrelvir/ritonavir following its prescription and concomitant use remdesivir and anti-SARS-CoV-2 monoclonal antibodies could introduce a bias. The intensity of the follow-up is also another possible bias because those who were prescribed the drug may have better medical supervision, which may also contribute to a lower risk of hospitalization.

Furthermore, the conditions of access to RT-PCR testing could result in those who had access to testing being more vulnerable individuals, thereby overestimating the risk of hospitalization in the control group. In addition, although we excluded individuals hospitalized at the index date, individuals who may have tested positive in the emergency department and spent more than one day were not excluded, which could overestimate the risk of hospitalization in the control group. These biases could lead to an underestimate of the real-life effectiveness of nirmatrelvir/ritonavir. However, given that access to nirmatrelvir/ritonavir is restricted to individuals at high risk of progression to severe disease, this bias may be minimized.

Another limitation is the lack of data regarding prior SARS-CoV-2 infections and hybrid immunity, a complicated variable due to the discontinuation of systematic RT-PCR tests in the province of Québec and the fact that antigenic self-test results were not reported in our databases. A study using hybrid immunity data, including the type of hybrid immunity with the number of previous infections and the duration since the last immunogenic event, would be of great interest and could guide decision-makers in fine-tuning vaccine recall policies. In addition, considering that a complete primary vaccination course is no longer up to date, the Comité sur l’immunisation du Québec [Québec immunization Committee] now considers that the primary vaccination with two doses followed by a first-booster dose constitutes the basic vaccination against COVID-19, and another booster campaign is ongoing for certain groups in the fall of 2022 to consolidate acquired immunity against COVID-19 ^61^.

Finally, there are limitations in the generalizability of the results of this study. Nirmatrelvir/ritonavir was prescribed only to those outpatients at high risk of severe COVID-19. In addition, since January 2022, RT-PCR tests have no longer been available for the entire general population in the province of Québec, and a selection bias may have been a possible influence on the criteria of accessibility to RT-PCR tests on the characteristics of the control group. Thus, treatment and control groups were not representative of the general population and the results may not be generalizable to the general population. It should also be noted that the advent of new COVID-19 variants may affect the real-life efficacy of nirmatrelvir/ritonavir and our results may no longer apply in this setting.

In summary, our large-scale province-wide cohort study suggests that treatment with nirmatrelvir/ritonavir is associated with a marked reduction of COVID-19-associated hospitalization in the era of Omicron, especially in unvaccinated infected individuals with high risk of severe COVID-19, those with an incomplete primary vaccination course, those that are severely immunocompromised individuals regardless of their vaccination and high-risk individuals aged 70 years and older with a completed primary vaccination course when six months or more has elapsed between infection and the last dose of the vaccine. Except for severely immunocompromised outpatients, no evidence of benefit was found in outpatients with a complete primary vaccination course whose last dose of the vaccine was given within six months.

## Data Availability

All data produced in the present work are contained in the manuscript.

## Acknowledgments

The authors thank their colleagues from INESSS, Michèle Paré and José Perez who contributed to creating the COVID-19 cohort which was used in this study. The authors would also like to thank Delphine Rochefort, Jim Boulanger, and Gino Boily, who helped in identifying individuals with severe immunosuppression and Jordie Croteau for his comments on the statistical methods. Members of the INESSS’s COVID-19 therapeutic task force for their comments on the method and results during the meeting in autumn 2022.

## REFERENCES

1. WHO. WHO Coronavirus (COVID-19) Dashboard. 2022; https://covid19.who.int/. Accessed 14 February 2023.

2. Hammond J, Leister-Tebbe H, Gardner A, et al. Oral Nirmatrelvir for High-Risk, Nonhospitalized Adults with Covid-19. N Engl J Med. 2022.

3. Mallapaty S. COVID-19: How Omicron overtook Delta in three charts. Nature. 2022.

4. Rubin EJ, Baden LR, Morrissey S. Audio Interview: Understanding the Omicron Variant of SARS-CoV-2. N Engl J Med. 2022;386(8):e27.

5. INESSS. NIRMATRELVIR / RITONAVIR (PAXLOVID). 2022; https://www.inesss.qc.ca/covid-19/traitements-specifiques-a-la-covid-19/nirmatrelvir-/-ritonavir-paxlovid.html. Accessed 7 novembre 2022.

6. Canadian Institute for Health Information. CIHI’s Population Grouping Methodology 1.3. 2021; https://www.cihi.ca/sites/default/files/document/cihi-population-grouping-methodology-v1_3-overview-outputs-manual-en.pdf. Accessed 28 novembre 2022.

7. Zou G. A modified poisson regression approach to prospective studies with binary data. Am J Epidemiol. 2004;159(7):702–706.

8. Furukawa TA, Guyatt GH, Griffith LE. Can we individualize the ‘number needed to treat’? An empirical study of summary effect measures in meta-analyses. Int J Epidemiol. 2002;31(1):72–76.

9. Bajema KL, Dahl RM, Evener SL, et al. Comparative Effectiveness and Antibody Responses to Moderna and Pfizer-BioNTech COVID-19 Vaccines among Hospitalized Veterans - Five Veterans Affairs Medical Centers, United States, February 1- September 30, 2021. MMWR Morb Mortal Wkly Rep. 2021;70(49):1700–1705.

10. Tenforde MW, Self WH, Naioti EA, et al. Sustained Effectiveness of Pfizer-BioNTech and Moderna Vaccines Against COVID-19 Associated Hospitalizations Among Adults - United States, March-July 2021. MMWR Morb Mortal Wkly Rep. 2021;70(34):1156–1162.

11. Auvigne V, Vaux S, Le Strat Y, et al. Severe hospital events following symptomatic infection with Sars-CoV-2 Omicron and Delta variants in France, December 2021 - January 2022: a retrospective, population-based, matched cohort study. medRxiv. 2022;04.

12. Bager P, Wohlfahrt J, Bhatt S, et al. Risk of hospitalisation associated with infection with SARS-CoV-2 omicron variant versus delta variant in Denmark: an observational cohort study. Lancet Infect Dis. 2022.

13. Sheikh A, Kerr S, Woolhouse M, McMenamin J, Robertson C, Collaborators EI. Severity of omicron variant of concern and effectiveness of vaccine boosters against symptomatic disease in Scotland (EAVE II): a national cohort study with nested test-negative design. Lancet Infect Dis. 2022.

14. Nyberg T, Ferguson NM, Nash SG, et al. Comparative analysis of the risks of hospitalisation and death associated with SARS-CoV-2 omicron (B.1.1.529) and delta (B.1.617.2) variants in England: a cohort study. Lancet. 2022.

15. Skarbinski J, Wood MS, Chervo TC, et al. Risk of severe clinical outcomes among persons with SARS-CoV-2 infection with differing levels of vaccination during widespread Omicron (B.1.1.529) and Delta (B.1.617.2) variant circulation in Northern California: A retrospective cohort study. Lancet Reg Health Am. 2022:100297.

16. Pfizer. Pfizer Reports Additional Data on PAXLOVID™ Supporting Upcoming New Drug Application Submission to U.S. FDA. 2022; https://www.pfizer.com/news/press-release/press-release-detail/pfizer-reports-additional-data-paxlovidtm-supporting. Accessed 19 août 2022.

17. Dryden-Peterson S, Kim A, Kim AY, et al. Nirmatrelvir plus ritonavir for early COVID-19 and hospitalization in a large US health system. medRxiv. 2022:2022.2006.2014.22276393.

18. Aggarwal NR, Molina KC, Beaty LE, Bennett TD, Carlson NE, Ginde AA. Real-world Use of Nirmatrelvir-Ritonavir in COVID-19 Outpatients During the Emergence of Omicron Variants BA.2/BA2.12.1. medRxiv. 2022:2022.2009.2012.22279866.

19. Arbel R, Wolff Sagy Y, Hoshen M, et al. Nirmatrelvir Use and Severe Covid-19 Outcomes during the Omicron Surge. N Engl J Med. 2022.

20. Brown PA, McGuinty M, Argyropoulos C, et al. Early experience with modified dose nirmatrelvir/ritonavir in dialysis patients with coronavirus disease-2019. medRxiv. 2022;21.

21. Devresse A, Sebastien B, De Greef J, et al. Safety, Efficacy, and Relapse of Nirmatrelvir-Ritonavir in Kidney Transplant Recipients Infected With SARS-CoV-2. Kidney International Reports. 2022.

22. Ganatra S, Dani SS, Ahmad J, et al. Oral Nirmatrelvir and Ritonavir in Non-hospitalized Vaccinated Patients with Covid-19. Clinical Infectious Diseases. 2022:ciac673.

23. Gentile I, Scotto R, Moriello NS, et al. Nirmatrelvir/ritonavir and molnuipiravir in the treatment of mild/moderate COVID-19: results of a real-life study. medRxiv. 2022:2022.2008.2023.22278585.

24. Hashash JG, Desai A, Kochhar GS, Farraye FA. Efficacy of Paxlovid and Lagevrio for COVID-19 Infection in patients with Inflammatory Bowel Disease: A Propensity Matched Study. Clin Gastroenterol Hepatol. 2022.

25. Hedvat J, Lange NW, Salerno DM, et al. COVID-19 therapeutics and outcomes among solid organ transplant recipients during the Omicron BA.1 era. Am J Transplant. 2022.

26. Lewnard JA, Malden D, Hong V, et al. Effectiveness of nirmatrelvir-ritonavir against hospital admission: a matched cohort study in a large US healthcare system. medRxiv. 2022:2022.2010.2002.22280623.

27. Malden DE, Hong V, Lewin BJ, et al. Hospitalization and Emergency Department Encounters for COVID-19 After Paxlovid Treatment - California, December 2021-May 2022. MMWR Morb Mortal Wkly Rep. 2022;71(25):830–833.

28. Najjar-Debbiny R, Gronich N, Weber G, et al. Effectiveness of Paxlovid in Reducing Severe COVID-19 and Mortality in High Risk Patients. Clin Infect Dis. 2022.

29. Razonable RR, O’Horo JC, Hanson SN, et al. Outcomes of Bebtelovimab Treatment is Comparable to Ritonavir-boosted Nirmatrelvir among High-Risk Patients with Coronavirus Disease-2019 during SARS-CoV-2 BA.2 Omicron Epoch. The Journal of Infectious Diseases. 2022:jiac346.

30. Salerno DM, Jennings DL, Lange NW, et al. Early clinical experience with nirmatrelvir/ritonavir for treatment of COVID-19 in solid organ transplant recipients. Am J Transplant. 2022.

31. Vora SB, Englund JA, Trehan I, et al. Monoclonal antibody and antiviral therapy for treatment of mild-to-moderate COVID-19 in pediatric patients. medRxiv. 2022:2022.2003.2016.22272511.

32. Wai AK, Chan CY, Cheung AW, et al. Association of Molnupiravir and Nirmatrelvir-Ritonavir with preventable mortality, hospital admissions and related avoidable healthcare system cost among high-risk patients with mild to moderate COVID-19. Lancet Reg Health West Pac. 2022:100602.

33. Wong CKH, Au ICH, Lau KTK, Lau EHY, Cowling BJ, Leung GM. Real-world effectiveness of early molnupiravir or nirmatrelvir-ritonavir in hospitalised patients with COVID-19 without supplemental oxygen requirement on admission during Hong Kong’s omicron BA.2 wave: a retrospective cohort study. Lancet Infect Dis. 2022.

34. Wong CKH, Au ICH, Lau KTK, Lau EHY, Cowling BJ, Leung GM. Real-world effectiveness of molnupiravir and nirmatrelvir plus ritonavir against mortality, hospitalisation, and in-hospital outcomes among community-dwelling, ambulatory patients with confirmed SARS-CoV-2 infection during the omicron wave in Hong Kong: an observational study. Lancet. 2022;400(10359):1213–1222.

35. Yip TCF, Lui GCY, Lai MSM, et al. Impact of the use of oral antiviral agents on the risk of hospitalization in community COVID-19 patients. Clin Infect Dis. 2022.

36. Zhou X, Kelly SP, Liang C, et al. Real-World Effectiveness of Nirmatrelvir/Ritonavir in Preventing Hospitalization Among Patients With COVID-19 at High Risk for Severe Disease in the United States: A Nationwide Population-Based Cohort Study. medRxiv. 2022:2022.2009.2013.22279908.

37. Qian G, Wang X, Patel NJ, et al. Outcomes with and without outpatient SARS-CoV-2 treatment for patients with COVID-19 and systemic autoimmune rheumatic diseases: A retrospective cohort study. medRxiv. 2022.

38. Schwartz KL, Wang J, Tadrous M, et al. Real-world effectiveness of nirmatrelvir/ritonavir use for COVID-19: A population-based cohort study in Ontario, Canada. medRxiv. 2022:2022.2011.2003.22281881.

39. Xie Y, Choi T, Al-Aly Z. Nirmatrelvir and the Risk of Post-Acute Sequelae of COVID-19. medRxiv. 2022:2022.2011.2003.22281783.

40. Shah. Paxlovid Associated with Decreased Hospitalization Rate Among Adults with COVID-19 — United States. MMWR Morb Mortal Wkly Rep. 2022.

41. Pfizer Canada. Monographie - ^Pr^PAXLOVID^MC^. 2022; https://www.pfizer.ca/sites/default/files/202201/PAXLOVID_PM_Fr_259186_17Jan2022.pdf. Accessed 17 janvier 2022.

42. Gottlieb RL, Vaca CE, Paredes R, et al. Early Remdesivir to Prevent Progression to Severe Covid-19 in Outpatients. N Engl J Med. 2021.

43. Gupta A, Gonzalez-Rojas Y, Juarez E, et al. Effect of Sotrovimab on Hospitalization or Death Among High-risk Patients With Mild to Moderate COVID-19: A Randomized Clinical Trial. Jama. 2022.

44. Jayk Bernal A, Gomes da Silva MM, Musungaie DB, et al. Molnupiravir for Oral Treatment of Covid-19 in Nonhospitalized Patients. N Engl J Med. 2021.

45. Montgomery H, Hobbs FDR, Padilla F, et al. Efficacy and safety of intramuscular administration of tixagevimab-cilgavimab for early outpatient treatment of COVID-19 (TACKLE): a phase 3, randomised, double-blind, placebo-controlled trial. Lancet Respir Med. 2022.

46. Anderson AS, Caubel P, Rusnak JM. Nirmatrelvir-Ritonavir and Viral Load Rebound in Covid-19. The New England journal of medicine. 2022;387(11):1047–1049.

47. Boucau J, Uddin R, Marino C, et al. Characterization of virologic rebound following nirmatrelvir-ritonavir treatment for COVID-19. Clin Infect Dis. 2022.

48. Carlin AF, Clark AE, Chaillon A, et al. Virologic and Immunologic Characterization of COVID-19 Recrudescence after Nirmatrelvir/Ritonavir Treatment. Res Sq. 2022.

49. Carlin AF, Clark AE, Chaillon A, et al. Virologic and Immunologic Characterization of COVID-19 Recrudescence after Nirmatrelvir/Ritonavir Treatment. Clin Infect Dis. 2022.

50. Charness ME, Gupta K, Stack G, et al. Rebound of SARS-CoV-2 Infection after Nirmatrelvir-Ritonavir Treatment. The New England journal of medicine. 2022;387(11):1045–1047.

51. Coulson JM, Adams A, Gray LA, Evans A. COVID-19 “Rebound” associated with nirmatrelvir/ritonavir pre-hospital therapy. J Infect. 2022.

52. Dai EY, Lee KA, Nathanson AB, et al. Viral Kinetics of Severe Acute Respiratory Syndrome Coronavirus 2 (SARS-CoV-2) Omicron Infection in mRNA-Vaccinated Individuals Treated and Not Treated with Nirmatrelvir-Ritonavir. medRxiv. 2022.

53. Epling BP, Rocco JM, Boswell KL, et al. COVID-19 redux: clinical, virologic, and immunologic evaluation of clinical rebound after nirmatrelvir/ritonavir. medRxiv. 2022:2022.2006.2016.22276392.

54. Epling BP, Rocco JM, Boswell KL, et al. Clinical, Virologic, and Immunologic Evaluation of Symptomatic Coronavirus Disease 2019 Rebound Following Nirmatrelvir/Ritonavir Treatment. Clin Infect Dis. 2022.

55. Gupta. Rapid Relapse of Symptomatic SARS-CoV-2 Infection Following Early Suppression with Nirmatrelvir/Ritonavir. Research Square. 2022.

56. Ranganath N, O’Horo JC, Challener DW, et al. Rebound Phenomenon after Nirmatrelvir/Ritonavir Treatment of Coronavirus Disease-2019 in High-Risk Persons. Clin Infect Dis. 2022.

57. Soares H, Baniecki ML, Cardin R, et al. Viral Load Rebound in Placebo and Nirmatrelvir-Ritonavir Treated COVID-19 Patients is not Associated with Recurrence of Severe Disease or Mutations. Research Square. 2022.

58. Wang L, Berger NA, Davis PB, Kaelber DC, Volkow ND, Xu R. COVID-19 rebound after Paxlovid and Molnupiravir during January-June 2022. medRxiv. 2022:2022.2006.2021.22276724.

59. Wang L, Volkow ND, Davis PB, Berger NA, Kaelber DC, Xu R. COVID-19 rebound after Paxlovid treatment during Omicron BA.5 vs BA.2.12.1 subvariant predominance period. medRxiv. 2022:2022.2008.2004.22278450.

60. Deo R, Choudhary MC, Moser C, et al. Viral and Symptom Rebound in Untreated COVID-19 Infection. medRxiv. 2022:2022.2008.2001.22278278.

61. Quebec Immunization Committee. Basic vaccination against COVID-19 and periodic consolidation of immunity. 2022; https://www.inspq.qc.ca/publications/3220-vaccination-covid-consolidation. Accessed 23 août 2022.

