## Supplemental Files for "REAL-WORLD EFFECTIVENESS OF NIRMATRELVIR/RITONAVIR ON COVID-19-ASSOCIATED HOSPITALIZATION PREVENTION: A POPULATION-BASED COHORT STUDY IN THE PROVINCE OF QUÉBEC, CANADA"

**Appendix A.** Sensitivity Analysis: Multivariable analysis using all sample without propensity score matching

**Appendix B.** Sensitivity Analysis for different definition of the outcome

**Appendix C.** Health Conditions

**Appendix D.** Identification of severe immunosuppression

**APPENDIX**

**Appendix A.** Sensitivity Analysis: Multivariable analysis using all sample without propensity score matching

**Table 1.** Risk of COVID-19-associated hospitalization among outpatients with high-risk of progression to severe COVID-19 who received nirmatrelvir/ritonavir prescription compared to controls

| **Population** | **Group** | **Population** | **Hospitalisations** | | **Univariable Poisson regression with robust error variance** | | | **Multivariable Poisson regression with robust error variance^1^** | | |
| --- | --- | --- | --- | --- | --- | --- | --- | --- | --- | --- |
|  |  | **N** | **N** | **%** | **RR** | **95% CI** | **P-value** | **RR** | **95% CI** | **P-value** |
| All (incomplete and complete primary vaccination) | Control | 242,337 | 8,293 | 3.4 | 1 |  |  | 1 |  |  |
|  | Treated | 16,601 | 356 | 2.1 | **0.63** | **0.56; 0.70** | **<0.001** | **0.15** | **0.13; 1.17** | **<0.001** |
| Incomplete primary vaccination | Control | 18,123 | 1,054 | 5.8 | 1 |  |  |  |  |  |
|  | Treated | 12,699 | 62 | 0.5 | **0.08** | **0.07; 0.11** | **<0.001** | **0.02** | **0.02; 0.03** | **<0.001** |
| Complete primary vaccination | Control | 224,214 | 7,239 | 3.2 | 1 |  |  | 1 |  |  |
|  | Treated | 3,902 | 294 | 7.5 | 2.33 | 2.09; 2.61 | <0.001 | 0.92 | 0.82; 1.03 | 0.134 |
| Complete primary vaccination with last vaccine dose ≤ 6 months | Control | 139,544 | 4,987 | 3.6 | 1 |  |  | 1 |  |  |
|  | Treated | 2,864 | 223 | 7.8 | 2.18 | 1.92; 2.48 | <0.001 | 1.0 | 0.88; 1.14 | 0.965 |
| Complete primary vaccination  With last vaccine dose > 6 months | Control | 84,670 | 2,252 | 2.7 | **1** |  |  | **1** |  |  |
|  | Treated | 1,038 | 71 | 6.8 | **2.57** | **2.05; 3.23** | **<0.001** | **0.71** | **0.56; 0.89** | **0.004** |

*1: Adjusted for age, sex, region of residence, number of vaccine dose, time since last vaccine dose, COVID-19 wave, number of health conditions, respiratory and cardiovascular condition, immunosuppression condition, cancer conditions.*

*Abbreviations: RR: Relative Risk; 95% CI: 95% Confidence Intervals.*

**Table 2.** Risk of COVID-19-associated hospitalization in outpatient with high-risk of progression to severe COVID-19 s who received nirmatrelvir/ritonavir prescription compared to controls in subgroup of outpatients with a complete primary vaccination

| **Population** | **Group** | **N personnes** | **N hospit.** | **% hospit.** | **Univariable Poisson regression with robust error variance** | | | **Multivariable Poisson regression with robust error variance^1^** | | |
| --- | --- | --- | --- | --- | --- | --- | --- | --- | --- | --- |
|  |  | **N** | **N** | **%** | **RR** | **95% CI** | **P-value** | **RR** | **95% CI** | **P-value** |
| *Less than 70 years (All)* | Control | 183,690 | 2,037 | 1.1 | 1 |  |  | 1 |  |  |
|  | Treated | 2,137 | 81 | 3.8 | 3.42 | 2.75; 4.25 | <0.001 | 1.15 | 0.91; 1.46 | 0.242 |
| *Less than 70 years (last dose ≤ 6 months)* | Control | 105,265 | 1,136 | 1.1 | 1 |  |  | 1 |  |  |
|  | Treated | 1,436 | 56 | 3.9 | 3.61 | 2.78; 4.70 | <0.001 | 1.15 | 0.87; 1.53 | 0.326 |
| Less than 70 years (last dose > 6 months) | Control | 78,425 | 901 | 1.2 | 1 |  |  |  |  |  |
|  | Treated | 701 | 25 | 3.6 | 3.10 | 2.10; 4.59 | <0.001 | 1.13 | 0.74; 1.72 | 0.574 |
| 70 years and older (All) | Control | 40,524 | 5,202 | 12.8 | 1 |  |  |  |  |  |
|  | Treated | 1,765 | 213 | 12.1 | 0.94 | 0.83; 1.07 | 0.346 | 0.82 | 0.72; 0.94 | 0.003 |
| *70 years and older (last dose ≤ 6 months)* | Control | 34,279 | 3,851 | 11.2 | 1 |  |  |  |  |  |
|  | Treated | 1,428 | 167 | 11.7 | 1.04 | 0.90; 1.20 | 0.589 | 0.93 | 0.80; 1.07 | 0.310 |
| 70 years and older (last dose > 6 months) | Control | 6,245 | 1,351 | 21.6 | 1 |  |  |  |  |  |
|  | Treated | 337 | 46 | 13.7 | 0.63 | 0.48; 0.83 | 0.001 | 0.58 | 0.44; 0.76 | <0.001 |
| Severely immunocompromised (All) | Control | 2,012 | 452 | 22.5 | 1 |  |  |  |  |  |
|  | Treated | 524 | 63 | 12.0 | 0.54 | 0.42; 0.68 | <0.001 | 0.60 | 0.47; 0.76 | <0.001 |
| Severely immunocompromised (last dose ≤ 6 months) | Control | 1,502 | 334 | 22.2 | 1 |  |  |  |  |  |
|  | Treated | 375 | 44 | 11.7 | 0.53 | 0.39; 0.71 | <0.001 | 0.60 | 0.45; 0.80 | 0.001 |
| Severely immunocompromised (last dose > 6 months) | Control | 510 | 118 | 23.1 | 1 |  |  |  |  |  |
|  | Treated | 149 | 19 | 12.8 | 0.55 | 0.35; 0.86 | 0.009 | 0.60 | 0.38; 0.93 | 0.023 |

*1: Adjusted for age, sex, region of residence, number of vaccine dose, time since last vaccine dose, COVID-19 wave, number of health conditions, respiratory and cardiovascular condition, immunosuppression condition, cancer conditions.*

*Abbreviations: RR: Relative Risk; 95% CI: 95% Confidence Intervals.*

**Appendix B.** Sensitivity Analysis for different definition of the outcome

**Table 1.** Analysis was performed using hospitalization due to COVID-19 as outcome, i.e., hospitalization with COVID-19 as the main cause of admission

**Table 2.** Analysis was performed using COVID-19 associated hospitalization and death as outcome, i.e., the outcome encompassed hospitalization and/or death associated with COVID-19 and occurring in the 30 days following the index date.

**Table 1.** Risk of hospitalization due to COVID-19 among outpatients with high-risk of progression to severe COVID-19 who received nirmatrelvir/ritonavir prescription compared to controls

|  |  | **Before Propensity Score Matching** | | | | | | **After Propensity Score Matching** | | | | | |
| --- | --- | --- | --- | --- | --- | --- | --- | --- | --- | --- | --- | --- | --- |
| **Population** | **Group** | **Population** | **Hospitalisations** | | **Unadjusted Poisson regression with robust error variance^1^** | | | **Population** | **Hospitalisations** | | **Adjusted Poisson regression with robust error variance^1^** | | |
|  |  | **N** | **N** | **%** | **RR** | **95% CI** | **P-value** | **N** | **N** | **%** | **RR** | **95% CI** | **P-value** |
| All (incomplete and complete primary vaccination) | Control | 238,850 | 4,806 | 2.0 | 1 |  |  | 8,087 | 635 | 7.9 | 1 |  |  |
|  | Treated | 16,469 | 224 | 1.4 | 0.68 | 0.59; 0.77 | <0.001 | 8,087 | 180 | 2.2 | 0.29 | 0.25; 0.34 | <0.001 |
| Incomplete primary vaccination | Control | 17,668 | 599 | 3.4 | 1 |  |  | 4,514 | 405 | 9.0 | 1 |  |  |
|  | Treated | 12,675 | 38 | 0.3 | 0.09 | 0.06; 0.12 | <0.001 | 4,514 | 11 | 0.2 | 0.03 | 0.02; 0.05 | <0.001 |
| Complete primary vaccination | Control | 221,182 | 4,207 | 1.9 | 1 |  |  | 3,551 | 214 | 6.0 | 1 |  |  |
|  | Treated | 3,794 | 186 | 4.9 | 2.58 | 2.23; 2.97 | <0.001 | 3,551 | 174 | 4.9 | 0.85 | 0.70; 1.02 | 0.081 |
| Complete primary vaccination with last vaccine dose ≤ 6 months | Control | 137,557 | 3,000 | 2.2 | 1 |  |  | 2,604 | 147 | 5.7 | 1 |  |  |
|  | Treated | 2,782 | 141 | 5.1 | 2.32 | 1.97; 2.74 | <0.001 | 2,604 | 134 | 5.2 | 0.93 | 0.75; 1.16 | 0.532 |
| Complete primary vaccination with last vaccine dose > 6 months | Control | 83,625 | 1,207 | 1.4 | 1 |  |  | 843 | 58 | 6.9 | 1 |  |  |
|  | Treated | 1,012 | 45 | 4.5 | 3.08 | 2.30; 4,12 | <0.001 | 843 | 34 | 4.0 | 0.61 | 0.41; 0.91 | 0.015 |
| Complete primary vaccination and aged less than 70 years (All) | Control | 182,508 | 855 | 0.5 | 1 |  |  | 1,829 | 29 | 1.6 | 1 |  |  |
|  | Treated | 2,105 | 49 | 2.3 | 4.97 | 3.74; 6.61 | <0.001 | 1,829 | 42 | 2.3 | 1.49 | 0.95; 2.34 | 0.084 |
| Complete primary vaccination and aged less than 70 years (last dose ≤ 6 months) | Control | 104,643 | 514 | 0.5 | 1 |  |  | 1,212 | 24 | 2.0 | 1 |  |  |
|  | Treated | 1,415 | 35 | 2.5 | 5.04 | 3.59; 7.06 | <0.001 | 1,212 | 31 | 2.6 | 1.31 | 0.78; 2.19 | 0.311 |
| Complete primary vaccination and aged less than 70 years (last dose > 6 months) | Control | 77,865 | 341 | 0.4 | 1 |  |  | 569 | 7 | 1.2 | 1 |  |  |
|  | Treated | 690 | 14 | 2.0 | 4.63 | 2.73; 7.86 | <0.001 | 569 | 9 | 1.6 | 1.49 | 0.58; 3.83 | 0.413 |
| Complete primary vaccination and aged 70 years and older (All) | Control | 38,674 | 3,352 | 8.7 | 1 |  |  | 1,586 | 165 | 10.4 | 1 |  |  |
|  | Treated | 1,689 | 137 | 8.1 | 0.94 | 0.79; 1.10 | 0.427 | 1,586 | 122 | 7.7 | 0.73 | 0.58; 0.91 | 0.005 |
| Complete primary vaccination and aged 70 years and older (last dose ≤ 6 months) | Control | 32,914 | 2,486 | 7.6 | 1 |  |  | 1,316 | 116 | 8.8 | 1 |  |  |
|  | Treated | 1,367 | 106 | 7.8 | 1.03 | 0.85; 1.24 | 0.783 | 1,316 | 97 | 7.4 | 0.83 | 0.64; 1.07 | 0.142 |
| Complete primary vaccination and aged 70 years and older (last dose > 6 months) | Control | 5,760 | 866 | 15.0 | 1 |  |  | 239 | 48 | 20.1 | 1 |  |  |
|  | Treated | 322 | 31 | 9.6 | 0.64 | 0.46; 0.90 | 0.010 | 239 | 18 | 7.5 | 0.36 | 0.22; 0.59 | <0.001 |
| Complete primary vaccination and severely immunocompromised (All) | Control | 1,889 | 329 | 17.4 | 1 |  |  | 488 | 92 | 18.9 | 1 |  |  |
|  | Treated | 496 | 35 | 7.1 | 0.41 | 0.29; 0.57 | <0.001 | 488 | 34 | 7.0 | 0.37 | 0.26; 0.53 | <0.001 |
| Complete primary vaccination and severely immunocompromised (last dose ≤ 6 months) | Control | 1,413 | 245 | 17.3 | 1 |  |  | 348 | 55 | 15.8 | 1 |  |  |
|  | Treated | 352 | 21 | 6.0 | 0.34 | 0.22; 0.53 | <0.001 | 348 | 20 | 5.8 | 0.38 | 0.23; 0.62 | <0.001 |
| Complete primary vaccination and severely immunocompromised (last dose > 6 months) | Control | 476 | 84 | 17.7 | 1 |  |  | 134 | 28 | 20.9 | 1 |  |  |
|  | Treated | 144 | 14 | 9.7 | 0.55 | 0.32; 0.94 | 0.029 | 134 | 14 | 10.5 | 0.52 | 0.30; 0.89 | 0.018 |

*1: Confounding variables used to create propensity score were included in the final regression model and comprised age, sex, region of residence, number of vaccine dose, time since last vaccine dose, COVID-19 wave, number of health conditions, respiratory and cardiovascular condition, immunosuppression condition, cancer conditions.*

*Abbreviations: RR: Relative Risk; 95% CI: 95% Confidence Intervals.*

**Table 2.** Risk of COVID-19-associated hospitalization and death among outpatients with high-risk of progression to severe COVID-19 who received nirmatrelvir/ritonavir prescription compared to controls

|  |  | **Before Propensity Score Matching** | | | | | | **After Propensity Score Matching** | | | | | |
| --- | --- | --- | --- | --- | --- | --- | --- | --- | --- | --- | --- | --- | --- |
| **Population** | **Group** | **Population** | **Hospitalization** | | **Unadjusted Poisson regression with robust error variance^1^** | | | **Population** | **Hospitalization** | | **Adjusted Poisson regression with robust error variance^1^** | | |
|  |  | **N** | **N** | **%** | **RR** | **95% CI** | **P-value** | **N** | **N** | **%** | **RR** | **95% CI** | **P-value** |
| All | Control | 242,337 | 8,479 | 3.5 | 1 |  |  | 8,402 | 987 | 11.8 | 1 |  |  |
|  | Treated | 16,601 | 367 | 2.2 | 0.63 | 0.57; 0.70 | <0.001 | 8,402 | 308 | 3.7 | 0.32 | 0.28; 0.36 | <0.001 |
| Incomplete primary vaccination | Control | 18,123 | 1,073 | 5.9 | 1 |  |  | 4,701 | 661 | 14.1 | 1 |  |  |
|  | Treated | 12,699 | 64 | 0.5 | 0.09 | 0.07; 0.11 | <0.001 | 4,701 | 23 | 0.5 | 0.04 | 0.02; 0.05 | <0.001 |
| Complete primary vaccination | Control | 224,214 | 7,406 | 3.3 | 1 |  |  | 3,665 | 318 | 8.7 | 1 |  |  |
|  | Treated | 3,902 | 303 | 7.8 | 2.35 | 2.11; 2.63 | <0.001 | 3,665 | 285 | 7.8 | 0.93 | 0.80; 1.08 | 0.338 |
| Complete primary vaccination Last vaccine dose ≤ 6 months | Control | 139,544 | 5,111 | 3.7 | 1 |  |  | 2,688 | 232 | 8.6 | 1 |  |  |
|  | Treated | 2,864 | 231 | 8.1 | 2.20 | 1.94; 2.50 | <0.001 | 2,688 | 221 | 8.2 | 0.98 | 0.83; 1.17 | 0.853 |
| Complete primary vaccination Last vaccine dose > 6 months | Control | 84,670 | 2,295 | 2.7 | 1 |  |  | 885 | 98 | 11.1 | 1 |  |  |
|  | Treated | 1,038 | 72 | 6.9 | 2.56 | 2.04; 3.21 | <0.001 | 885 | 61 | 6.9 | 0.62 | 0.47; 0.83 | 0.001 |
| Complete primary vaccination Less than 70 years (All) | Control | 183,690 | 2,055 | 1.1 | 1 |  |  | 1,869 | 64 | 3.4 | 1 |  |  |
|  | Treated | 2,137 | 83 | 3.9 | 3.47 | 2.80; 4.31 | <0.001 | 1,869 | 76 | 4.1 | 1.21 | 0.88; 1.66 | 0.238 |
| Complete primary vaccination Less than 70 years (last dose ≤ 6 months) | Control | 105,265 | 1,147 | 1.1 | 1 |  |  | 1,231 | 40 | 3.3 | 1 |  |  |
|  | Treated | 1,436 | 58 | 4.0 | 3.71 | 2.86; 4.80 | <0.001 | 1,231 | 50 | 4.1 | 1.26 | 0.84; 1.88 | 0.261 |
| Complete primary vaccination Less than 70 years (last dose > 6 months) | Control | 78,425 | 908 | 1.2 | 1 |  |  | 579 | 17 | 2.9 | 1 |  |  |
|  | Treated | 701 | 25 | 3.6 | 3.08 | 2.08; 4.55 | <0.001 | 579 | 20 | 3.5 | 1.22 | 0.65; 2.30 | 0.534 |
| Complete primary vaccination 70 years and older (All) | Control | 40,524 | 5,351 | 13.2 | 1 |  |  | 1,678 | 268 | 16.0 | 1 |  |  |
|  | Treated | 1,765 | 220 | 12.5 | 0.94 | 0.83; 1.07 | 0.370 | 1,678 | 206 | 12.3 | 0.75 | 0.64; 0.89 | 0.001 |
| Complete primary vaccination 70 years and older (last dose ≤ 6 months) | Control | 34,279 | 3,964 | 11.6 | 1 |  |  | 1,388 | 187 | 13.5 | 1 |  |  |
|  | Treated | 1,428 | 173 | 12.1 | 1.05 | 0.91; 1.21 | 0.523 | 1,388 | 167 | 12.0 | 0.89 | 0.74; 1.08 | 0.249 |
| Complete primary vaccination 70 years and older (last dose > 6 months) | Control | 6,245 | 1,387 | 22.2 | 1 |  |  | 253 | 59 | 23.3 | 1 |  |  |
|  | Treated | 337 | 47 | 14.0 | 0.63 | 0.48; 0.82 | 0.001 | 253 | 31 | 12.3 | 0.51 | 0.35; 0.75 | 0.001 |
| Complete primary vaccination Immunocompromised (All) | Control | 2,012 | 468 | 23.3 | 1 |  |  | 519 | 97 | 18.7 | 1 |  |  |
|  | Treated | 524 | 64 | 12.2 | 0.53 | 0.41; 0.67 | <0.001 | 519 | 63 | 12.1 | 0.66 | 0.50; 0.88 | 0.005 |
| Complete primary vaccination and severely immunocompromised (last dose ≤ 6 months) | Control | 1,502 | 348 | 23.2 | 1 |  |  | 369 | 80 | 21.7 | 1 |  |  |
|  | Treated | 375 | 45 | 12.0 | 0.52 | 0.39; 0.69 | <0.001 | 369 | 44 | 11.9 | 0.58 | 0.42; 0.80 | 0.001 |
| Complete primary vaccination and severely immunocompromised (last dose > 6 months) | Control | 510 | 120 | 23.5 | 1 |  |  | 139 | 32 | 23.0 | 1 |  |  |
|  | Treated | 149 | 19 | 12.8 | 0.54 | 0.35; 0.85 | 0.007 | 139 | 19 | 13.7 | 0.61 | 0.37; 0.99 | 0.047 |

*1: Confounding variables used to create propensity score were included in the final regression model and comprised age, sex, region of residence, number of vaccine dose, time since last vaccine dose, COVID-19 wave, number of health conditions, respiratory and cardiovascular condition, immunosuppression condition, cancer conditions.*

**Appendix C.** Health Conditions

**Table 1.** List of CIHI Grouper codes used to identify targeted Health Conditions

| **RESPIRATORY, CARDIOVASCULAR AND OTHER CONDITIONS** | |
| --- | --- |
| **Pop Grouper Code** | **Condition Name** |
| A41 | Stroke |
| A43 | Transient ischemic attack |
| D02 | Congenital disorder of the respiratory system |
| D03 | Chronic obstructive pulmonary disease |
| D04 | Pulmonary hypertension |
| D05 | Other chronic lung disease |
| D06 | Asthma |
| D41 | Respiratory failure |
| E01 | Heart failure |
| E02 | Malformation of the cardiovascular system |
| E03 | Cardiac valve disease |
| E04 | Coronary artery disease |
| E05 | Arrhythmia |
| E06 | Other heart disease |
| E07 | Peripheral venous disease/ phlebitis/thrombophlebitis/DVT |
| E09 | Other vascular system disease |
| E10 | Hypertension |
| E11 | Aortic aneurysm |
| E12 | Peripheral artery disease |
| E41 | Acute myocardial infarction/shock/arrest |
| E43 | Unstable angina |
| G02 | Chronic liver disease (incl. hepatitis) |
| J09 | Hypercholesterolaemia and other dyslipidemia |
| J10 | Obesity |
| K01 | Chronic kidney disease/failure |
| J02 | Diabetes mellitus |
| **IMMUNOSUPPRESSIVE CONDITIONS** | |
| **Pop Grouper Code** | **Condition Name** |
| F06 | Inflammatory bowel (incl. crohn's, ulcerative colitis) |
| H01 | Rheumatoid & other inflammatory arthropathy (excl. gout) |
| H07 | Systemic connective tissue disorder (incl. lupus, scleroderma) |
| I01 | Autoimmune skin disorder |
| I02 | Papulosquamous disorder/ psoriasis |
| O01 | Disease of white blood cells (incl. neutropenia) |
| O03 | Coagulation & hemorrhagic disorder |
| O05 | Lymphatic system disorder (excl. spleen) |
| O06 | Disorder of immune mechanism |
| O07 | Other disease of blood & blood forming organs |
| P01 | Human immunodeficiency virus (HIV) infection |
| S03 | Transplant recipient |
| S41 | Transplant complication |
| A05 | Multiple sclerosis & other demyelinating disease of cns |
| **CANCER CONDITIONS** | |
| **Pop Grouper Code** | **Condition Name** |
| R01a | Brain cancer |
| R02c | Oral/ear/nose/throat cancer |
| R03d | Lung cancer |
| R04f | Colorectal cancer |
| R05f | Other digestive & hepatobiliary cancer |
| R06h | Musculoskeletal cancer |
| R07k | Renal cancer |
| R08i | Breast cancer |
| R09i | Skin cancer |
| R10j | Thyroid cancer |
| R11k | Prostate cancer |
| R12k | Bladder cancer |
| R13l | Ovarian cancer |
| R14l | Uterine cancer |
| R15l | Cervical cancer |
| R16o | Leukemia/lymphoma |
| R17 | Other & unspecified primary cancer |
| R18 | Metastatic cancer |

**Appendix D.** Identification of severe immunosuppression

**Table 1.** List of immunosuppressive cancers

| **Cancer group** | **ICD** | **ICD Code** | **Name** |
| --- | --- | --- | --- |
| Lymphoblastic Leukaemia | ICD-10-CA | C91 | Lymphoblastic Leukaemia |
|  | ICD-9 | 204 | Lymphoblastic Leukaemia |
| Myeloid Leukemia | ICD-10-CA | C92 | Myeloblastic Leukaemia |
|  | ICD-10-CA | C93 | Monocytic Leukaemia |
|  | ICD-10-CA | C940 | Acute Erythroid Leukaemia |
|  | ICD-10-CA | C941 | Chronic Erythremia |
|  | ICD-10-CA | C942 | Acute Megakaryoblastic Leukaemia |
|  | ICD-10-CA | C944 | Acute Panmyelosis With Myelofibrosis |
|  | ICD-10-CA | C945 | Acute Myelofibrosis |
|  | ICD-9 | 205 | Myeloid Leukemia |
|  | ICD-9 | 206 | Monocytic Leukemia |
|  | ICD-9 | 2070 | Erythroleukemia |
|  | ICD-9 | 2071 | Chronic Erythremia |
|  | ICD-9 | 2072 | Megakaryocytic Leukemia |
| Hodgkin Lymphoma | ICD-10-CA | C81 | Hodgkin Lymphoma |
|  | ICD-9 | 201 | Hodgkin's Disease |
| Non-Hodgkin Lymphoma | ICD-10-CA | C82 | Follicular Lymphoma |
|  | ICD-10-CA | C83 | Non-Follicular Lymphoma |
|  | ICD-10-CA | C84 | Mature T/NK-Cell Lymphomas |
|  | ICD-10-CA | C86 | Other Specified Types Of T/NK-Cell Lymphoma |
|  | ICD-10-CA | C85 | Other And Unspecified Types Of Non-Hodgkin Lymphoma |
|  | ICD-10-CA | C8808 | Other Lymphoplasmacytic Lymphoma |
|  | ICD-10-CA | C884 | Extranodal Marginal Zone B-Cell Lymphoma Of Mucosa-Associated Lymphoid Tissue [Maltlymphoma] |
|  | ICD-10-CA | C963 | True Histiocytic Lymphoma |
|  | ICD-9 | 200 | Lymphosarcoma And Reticulosarcoma |
|  | ICD-9 | 2020 | Nodular Lymphoma |
|  | ICD-9 | 2021 | Mycosis Fungoides |
|  | ICD-9 | 2028 | Other Malignant Lymphomas |
|  | ICD-9 | 2029 | Other And Unspecified Malignant Neoplasms Of Lymphoid And Histiocytic Tissue |
| Other Leukaemia | ICD-10-CA | C95 | Leukaemia Of Unspecified Cell Type |
|  | ICD-10-CA | C943 | Mast Cell Leukaemia |
|  | ICD-10-CA | C947 | Other Specified Leukaemias |
|  | ICD-10-CA | C901 | Plasma Cell Leukaemia |
|  | ICD-9 | 2024 | Leukemic Reticuloendotheliosis |
|  | ICD-9 | 2031 | Plasma Cell Leukemia |
|  | ICD-9 | 2078 | Other Specified Leukemia |
|  | ICD-9 | 208 | Leukemia Of Unspecified Cell Type |
| Multiple Myeloma | ICD-10-CA | C900 | Multiple Myeloma |
|  | ICD-10-CA | C902 | Extramedullary Plasmacytoma |
|  | ICD-10-CA | C903 | Solitary Plasmacytoma |
|  | ICD-9 | 203 | Multiple Myeloma And Immunoproliferative Neoplasms |
|  | ICD-9 | 2030 | Multiple Myeloma Except 2386 |
| Other Hematological Cancer | ICD-10-CA | C880 | Waldenström Macroglobulinaemia |
|  | ICD-10-CA | C881 | Alpha Heavy Chain Disease |
|  | ICD-10-CA | C882 | Other Heavy Chain Disease |
|  | ICD-10-CA | C883 | Immunoproliferative Small Intestinal Disease |
|  | ICD-10-CA | C887 | Other Malignant Immunoproliferative Diseases |
|  | ICD-10-CA | C889 | Malignant Immunoproliferative Disease, Unspecified |
|  | ICD-10-CA | C903 | Solitary Plasmacytoma |
|  | ICD-10-CA | C946 | Myelodysplastic And Myeloproliferative Disease, Not Elsewhere Classified |
|  | ICD-10-CA | C960 | Multifocal And Multisystemic (Disseminated) Langerhans-Cell Histiocytosis [Letterer-Siwe  Disease] |
|  | ICD-10-CA | C961 | Malignant Histiocytosis |
|  | ICD-10-CA | C962 | Malignant Mast Cell Tumour |
|  | ICD-10-CA | C964 | Sarcoma Of Dendritic Cells (Accessory Cells) |
|  | ICD-10-CA | C965 | Multifocal And Unisystemic Langerhans-Cell Histiocytosis |
|  | ICD-10-CA | C966 | Unifocal Langerhans-Cell Histiocytosis |
|  | ICD-10-CA | C967 | Other Specified Malignant Neoplasms of Lymphoid, Haematopoietic And Related Tissue |
|  | ICD-10-CA | C968 | Histiocytic Sarcoma |
|  | ICD-10-CA | C969 | Malignant Neoplasm of Lymphoid, Haematopoietic And Related Tissue, Unspecified |
|  | ICD-9 | 2022 | Sezary's Disease |
|  | ICD-9 | 2023 | Malignant Histiocytosis |
|  | ICD-9 | 2025 | Letterer-Siwe Disease |
|  | ICD-9 | 2026 | Malignant Mast Cell Tumors Except 2078 |
|  | ICD-9 | 2031 | Plasma Cell Leukemia |
|  | ICD-9 | 2038 | Other Immunoproliferative Neoplasms |
|  | ICD-9 | 2733 | Macroglobulinemia |

**Table 2.** List of immunosuppressive drugs

| **Therapy** | **Drugs** |
| --- | --- |
| Solid Organ Transplant + Immunosuppressive Drug | Azathioprine |
|  | Cyclosporine |
|  | Mycofenolate Mofetil |
|  | Mycophenolate Sodium |
|  | Sirolimus |
|  | Tacrolimus |
| Anti-B cell therapy   - Monoclonal antibodies targeting CD19, CD20, CD22, CD30 and BAFF | Ocrelizumab |
|  | Rituximab |
|  | Ofatumumab |
|  | Alemtuzumab |
| Alkylating agents for Rheumatoid Arthritis | Cyclophosphamide |
| Systemic Corticosteroids | Prednisone  (dose > 20 mg /day for at least 21 days) |
|  | Cortisone  (dose > 100 mg/day for at least 21 days) |
|  | Dexamethasone  (dose > 3 mg /day for at least 21 days) |
|  | Hydrocortisone  (dose > 80 mg/day for at least 21 days) |
